# The effect of the genetic liability to autism spectrum disorder on emotion recognition in young unaffected probands from a population-based cohort

**DOI:** 10.1101/19001230

**Authors:** Frank R Wendt, Carolina Muniz Carvalho, Joel Gelernter, Renato Polimanti

**Author notes:** Corresponding author: Renato Polimanti, PhD. Yale University School of Medicine, Department of Psychiatry. VA CT 116A2, 950 Campbell Avenue, West Haven, CT 06516, USA.

## Abstract

We investigated how ASD genetic risk relates to neurodevelopmental features (491 traits tested) via polygenic risk scoring (PRS) in 4,309 young non-ASD probands from the Philadelphia Neurodevelopmental Cohort. ASD PRS most strongly associated with the ability to correctly identify angry facial emotions in youths aged 11-17 years (R^2^=1.06%, p=1.38×10^−7^) and replicated similarly in older probands (>18 years) (R^2^=0.55%, p=0.036). The association in 11- to-17-year-old probands was independent of other psychiatric disorders, brain imaging phenotypes, and educational attainment. ASD PRS also associated with proband-reported emotionality and connectedness with others. The proband-reported irritability trait was highly correlated with angry facial emotion recognition (r^2^=0.159, p=2.74×10^−5^) but was independently associated with ASD PRS (R^2^=1.20%, p=1.18×10^−4^). Several informant-reported (i.e., typically mother-reported) traits were predicted by the proband’s ASD PRS, including duration of fear (R^2^=0.156%, p=0.001). These data indicate how genetic liability to ASD may influence neurodevelopment in the general population, especially the development of emotional intelligence.

## Introduction

Autism spectrum disorder (ASD) describes a group of pervasive neurodevelopmental disorders characterized by impaired social and communication skills. ASD typically manifests as a heterogeneous combination of repetitive and restrictive behavioral symptoms along with intellectual capabilities ranging from above average intelligence quotient (IQ) to intellectual disability.^1^ ASD affects approximately 1-1.5% of the general population and is diagnosed more frequently in males than females.^2^

ASD often causes serious impairment for affected individuals, although there is a large range and some function quite normally. Still, we need to resolve a puzzle on the population level – the polygenic risk that leads to ASD is composed of the effects of many risk loci, which are individually very small. What are the other effects of these risk loci? Is there any benefit to risk-allele carries who do not have enough collative risk to express ASD? Why are they maintained in the population? One possible explanation is that ASD PRS correlates with selection for better cognitive function.^3^ While that is a tantalizing clue, we might suspect that effects of ASD risk alleles in non-affected individuals might relate not just to cognitive function, but to traits more closely related to those that are seen in the pathological range in ASD subjects, such as those related to social-emotional reciprocity and nonverbal communication, difficulty developing and understanding relationships, repetitive motions, and hyper- or hypo-reactivity to sensory input. Investigating these relationships is the main premise of this investigation.

Using common genetic variation, Grove, *et al*.^4^ reported an estimated SNP-based observed-scale ASD heritability of ∼12% in Europeans. Several classes of genetic variant contribute to ASD liability, including de novo mutations (∼3%), non-additive genetic variation (∼4%), rare inherited variation (∼3%), and common inherited variation (∼49%).^5^ Furthermore, there were robust genetic and phenotype correlations between ASD, cognitive ability, educational attainment, and several behavioral traits.^4^ The highly polygenic nature and relatively high contribution of common genetic variation to ASD allude to a high degree of pleiotropy (i.e., a single genetic variant contributes small effects to several phenotypes or disorders) between the genetic liability to ASD, cognitive traits, and behavioral phenotypes. While these pleiotropic mechanisms have been investigated broadly, there is a paucity of data investigating the shared genetic information between specific neuropsychiatric domains and ASD.

We investigated the relationship between ASD risk alleles and hundreds of neuropsychiatric phenotypes in young probands from the Philadelphia Neurodevelopmental Cohort (PNC).^6-9^ Using polygenic risk scoring, we observed a significant positive relationship between genetic liability to ASD and (1) the ability to recognize anger and (2) the change in emotionality and connectedness with others. Conversely, we report a significant negative relationship between the genetic liability to ASD and the ability to correctly distinguish the age of others. These results highlight several features of human neurodevelopment in the young, such as emotional intelligence, as key pathophysiological targets for ASD etiology.

## Results

### Neuropsychiatric Trait Prediction

The genetic liability for ASD was used to predict 491 neuropsychiatric phenotypes in children not affected by ASD considering three age groups (young proband, N=1,035, age 8-10; middle probands, N=2,499, age 11-17; and adult probands, N=775 age ≥18) of the PNC (Figure 1, Table S1).^10^

**Figure 1.**
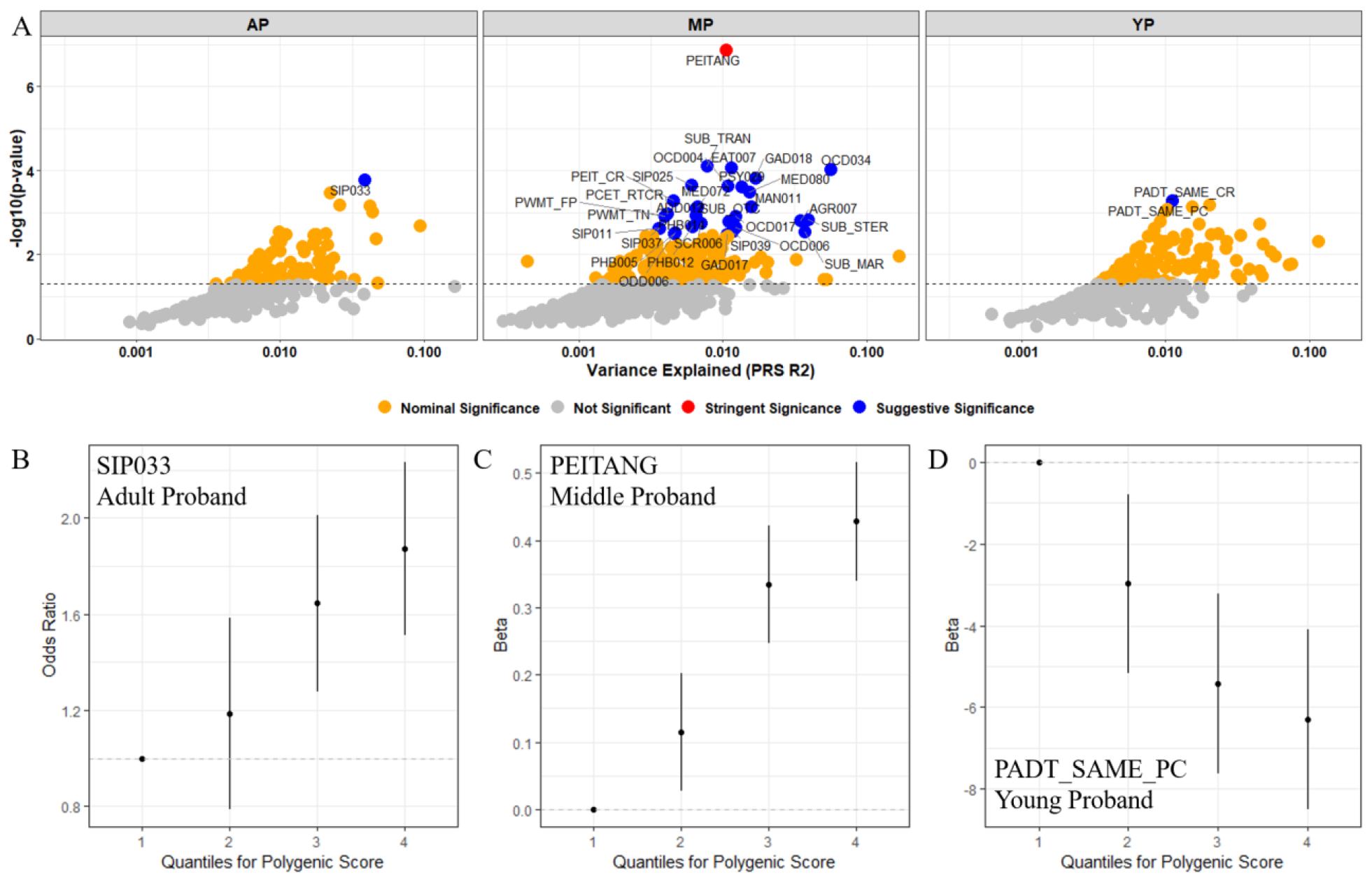
Overview of best-fit models for autism spectrum disorder (ASD) predicting neuropsychiatric phenotypes in adult (AP), middle (MP), and young (YP) probands of the Philadelphia Neurodevelopmental Cohort. (A) Maximum phenotypic variance explained (R^2^) by the best model fit for each trait given genetic liability to ASD. The dashed horizontal line represents the nominal significance threshold. (B-D) The relationship between binned ASD polygenic risk scores for AP, MP, and YP participants and the most significant phenotype predicted by ASD genetic liability: (B) *SIP033 Structural Interview for Prodromal Symptoms: “Has anyone pointed out to you that you are less emotional or connected to people than you used to be?”*; (C) *PEITANG: Number of correct responses to anger trials during completion of The Penn Emotional Identification Test (PEIT) for recognizing angry emotions*; (D) *PADT_SAME_PC: Percent of correct responses to test trials with no age difference during completion of the Penn Age Differentiation Test for detecting which face in a face pair appears older*. Note the lowest quartile in figures B-D represents the referent.

Polygenic risk for ASD significantly predicted the emotional intelligence phenotype *PEITANG* (Penn Emotional Identification Test (PEIT) recognition of angry facial emotions) (z-score = 5.28, R^2^ = 1.06%, p = 1.38×10^−7^; Figure 1) after applying a false discovery rate multiple-testing correction accounting for the number of phenotypes and PRS tested (N = 2,946,000 tests; ASD PRS → *PEITANG* FDR q = 6.81×10^−5^) in subjects aged 11-17 years. The ability of ASD PRS to predict *PEITANG* in the YP group nominally replicated with adult probands of mean age 19 ± 1.2 years (z-score = 2.10, R^2^ = 0.55%, p = 0.036). Combining the three age-stratified samples, a stronger association of ASD PRS with *PEITANG* phenotype was observed (z-score =5.69, R^2^ = 0.70%, p = 1.37×10^−8^). When binned by quartiles, there was a positive relationship between increased polygenic risk for ASD and the ability to recognize angry faces where the highest quartile had a 53% increase in the recognition of angry facial emotions when compared with lowest quartile of the ASD PRS distribution (beta_q4vs.q1_ = 0.427, p_q4vs.q1_ = 1.42×10^−6^; Figure 1). The *PEITANG* trait was nominally significantly correlated with all other emotion recognition traits and (0.007 ≤ r^2^ ≤ 0.378, 2.20×10^−16^ ≤ p ≤ 1.86×10^−5^) and four PEIT trial reaction times (happy, sad, anger, and fear trial reaction times; 0.001 ≤ r^2^ ≤ 0.007, 2.87×10^−8^ ≤ p ≤ 0.038; Table S2). To evaluate the relationship between ASD PRS and other PEIT emotion recognition tasks independently from the most significant association observed, *PEITANG* was included as a covariate in the model. After covarying for the effects of *PEITANG*, ASD PRS significantly predicted the ability to recognize two other facial emotions in PEIT: happiness (z-score = −2.41, R^2^ = 0.221%, p = 0.016) and neutral emotions (z-score = −2.05, R^2^ = 0.159%, p = 0.041; Figure 2). Furthermore, ASD PRS predicted *PEITANG* test reaction time (z-score = −2.08, R^2^ = 0.138%, p = 0.038) and reaction time for happy facial emotion trials after covarying for the effects of *PEITANG* (z-score = 2.21, R^2^ = 0.356%, p = 0.027; Figure 2). Conversely, the ability to decide which of two faces displays a more severe emotion (as measured by The Penn Emotion Differentiation Test (PEDT)) was poorly predicted by ASD polygenic risk (Figure S1).

**Figure 2.**
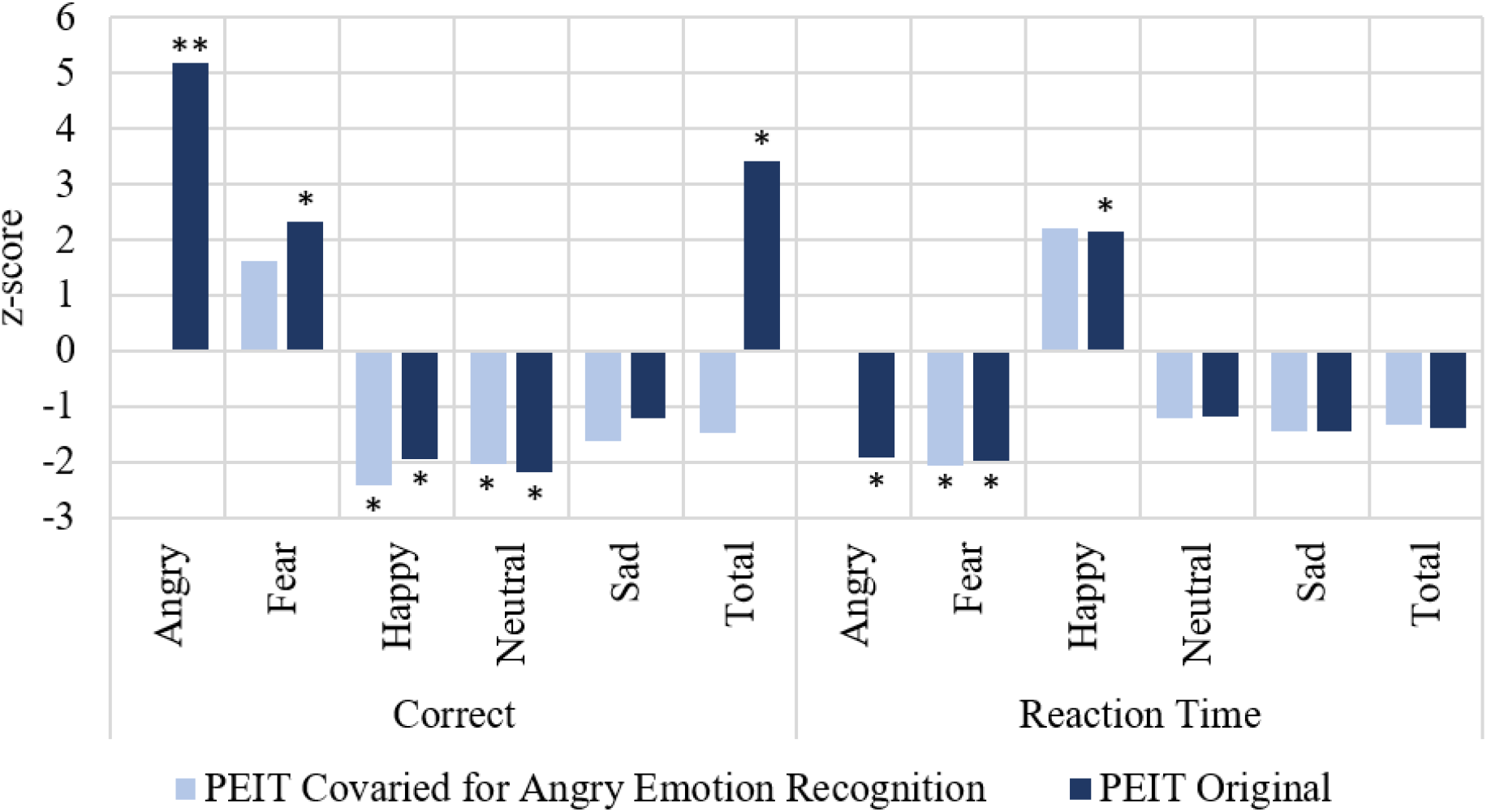
Prediction of facial emotion capabilities of the middle proband group (ages 11 to 17) of the Philadelphia Neurodevelopmental Cohort using polygenic risk (PRS) for autism spectrum disorder. The x-axis shows correctness and response time for each facial emotion using two neurocognitive instruments: The Penn Emotion Identification Test (shaded colors: original results; tinted colors: results covaried for the effects of *PEITANG*). Significance is indicated by * for p < 0.05 and ** for FDR Q < 0.05.

Considering a suggestive threshold based on a false discovery rate correction accounting for the number of phenotypes tested only (N = 491, FDR Q < 0.05), there were one, 30, and two additional phenotypes predicted by genetic liability to ASD in the adult, middle, and young proband groups, respectively (Table 2). The PNC traits *SIP033 (Structural Interview for Prodromal Symptoms: “Has anyone pointed out to you that you are less emotional or connected to people than you used to be?”)* and *PADT_SAME_PC (Penn Age Differentiation Test: percent of correct responses to trials with no age difference (60 total face pairs))* were predicted by ASD polygenic risk in the adult (z-score = 3.76, R^2^ = 3.84%, p = 1.69×10^−4^) and young (z-score = - 3.41, R^2^ = 1.06%, p = 5.08×10^−04^) probands, respectively (Figure 1). When binned by quartiles, there was a 2.39-fold increase between highest quartile and lowest quartile of the ASD PRS distribution in the odds of being told that you are less emotional/connected to people than previously in the adult proband group (beta_q4vs.q1_ = 0.873, p_q4vs.q1_ = 0.015; Figure 1B) and a 2.22-fold reduction in the percentage of total correct responses to age differentiation trials when no age difference was present in the young proband group (beta_q4vs.q1_ = 0.799, p_q4vs.q1_ = 0.004; Figure 1D).

**Table 1.** Samples sizes for each Philadelphia Neurodevelopmental Cohort proband group after quality control and selection of unrelated European samples.

| ***Proband*** | ***Age, years*** | ***Male N*** | ***Female N*** | ***Total N*** | ***Phenotype N*** |
| --- | --- | --- | --- | --- | --- |
| Young (YP) | 8-10 | 579 | 456 | 1,035 | 311 |
| Middle (MP) | 11-17 | 1,197 | 1,302 | 2,499 | 490 |
| Adult (AP) | 18 and over | 345 | 430 | 775 | 324 |

**Table 2.** Best model fit PRS of psychiatric disorder, educational attainment, and brain image-derived phenotype polygenic risk scores predicting the *PEITANG* phenotype in the Philadelphia Neurodevelopmental Cohort middle proband group (aged 11-17 years old).

| *Trait* | *Unadjusted* |  |  | *Adjusted for ASD PRS* |  |  |
| --- | --- | --- | --- | --- | --- | --- |
|  | *R2 (%)* | *z-score* | *p* | *R2 (%)* | *z-score* | *p* |
| Schizophrenia | 1.50 | 6.32 | $3.12 \times 10^{-10}$ | 0.898 | 4.89 | $1.05 \times 10^{-06}$ |
| Attention deficit hyperactivity disorder | 0.204 | 2.32 | 0.021 | 0.040 | 1.03 | 0.303 |
| Educational attainment | 0.463 | 3.49 | $4.98 \times 10^{-04}$ | 0.233 | 2.48 | 0.013 |
| Left cerebellum white matter volume | 0.580 | 3.91 | $9.62 \times 10^{-05}$ | 0.318 | 2.90 | $3.35 \times 10^{-04}$ |
| Mean diffusion tensor mode (MO) in sagittal stratum (left) on FA skeleton (from dMRI data) | 0.469 | -3.51 | $4.59 \times 10^{-04}$ | 0.333 | -2.97 | 0.003 |
| Mean L1 in anterior limb of internal capsule (right) on FA skeleton (from dMRI data) | 0.236 | -2.48 | 0.013 | 0.167 | -2.10 | 0.037 |
| Mean L3 in uncinate fasciculus (left) on FA skeleton (from dMRI data) | 0.286 | 2.74 | 0.006 | 0.269 | 2.67 | 0.008 |
| Right hemisphere central sulcus area (from Destrieux Atlas) | 0.560 | -3.84 | $1.27 \times 10^{-04}$ | 0.428 | -3.37 | $7.64 \times 10^{-04}$ |
| Left hemisphere cuneus gyrus thickness (from Destrieux Atlas) | 0.413 | 3.29 | 0.001 | 0.322 | 2.92 | 0.004 |

Eight out of 30 suggestively significant phenotypes from the middle proband group were nominally significantly correlated with *PEITANG* (0.002 ≤ r^2^ ≤ 0.378, 2.20×10^−16^ ≤ p ≤ 0.037, Table S3). To verify that predicting these phenotypes using ASD PRS was independent of the effects of *PEITANG, PEITANG* PRS was included as a covariate in each model. All 30 suggestively significant phenotypes from the middle proband group were significantly predicted by ASD after covarying for *PEITANG*. With the exception of *SIP011*, these additional suggestive relationships with ASD PRS had no change in effect after covarying for *PEITANG* (Figure S2). The phenotype *SIP011: SIPS-PRIME SCREEN-REVISED Structured Interview for Prodromal Symptoms: I think I might feel like my mind is “playing tricks” on me* was the only phenotype demonstrating a significant increase in effect coefficient after covarying for *PEITANG* (i.e., after covarying *PEITANG*, ASD PRS more strongly predicting *SIP011*; covaried z-score = 5.06, R^2^ = 0.948%, p = 6.98×10^−7^; original z-score = 3.05, R^2^ = 0.358%, p = 0.002; z-score_difference_ = −2.37 p_difference_ = 0.020).

### Influence of Related Brain Traits

To identify whether the association of the strongest genetically-predicted neuropsychiatric phenotype (i.e., *PEITANG*) was attributable to the shared genetic liability of ASD with other brain-related phenotypes rather than ASD *per se*, we next investigated the ability of polygenic liability to brain imaging phenotypes,^11^ other psychiatric disorders (i.e., attention deficit hyperactivity disorder (ADHD),^12^ anorexia nervosa,^13^ bipolar disorder,^14^ major depressive disorder,^15^ Tourette syndrome,^16^ obsessive compulsive disorder,^17^ and schizophrenia^18^), and educational attainment^19^ to predict *PEITANG* (Table 2 and Table S4). Among those traits, we selected fourteen brain imaging phenotypes and four psychiatric disorders that were significantly genetically correlated with ASD and included as covariates in evaluating the relationship between ASD and *PEITANG*.^20^ To remove genetically redundant traits, covariates were selected from genetically correlated trait pairs based on highest SNP-based heritability z-score (Figure 3) resulting in a total of six brain imaging phenotypes (left cerebellar white matter volume, mean diffusion tensor mode (MO) in sagittal stratum, mean L1 (i.e., strength of diffusion along the L1 principal axis of diffusion magnetic resonance imaging (dMRI)) in anterior limb of internal capsule (right), mean L3 (i.e., strength of diffusion along the L1 principal axis of dMRI) in uncinate fasciculus (left), right hemisphere central sulcus area, Left hemisphere cuneus gyrus thickness; for complete description of image acquisition and brain mapping, refer to Miller, *et al*. 2016^21^ and Elliot, *et al*. 2018^11^), two psychiatric disorders (attention deficit hyperactivity disorder and schizophrenia), and education attainment. After covarying, the polygenic risk for ASD significantly predicted the ability to recognize angry facial emotions in the middle proband group, considering both one covariate at a time and including all covariates in the model (Figure 4). The effects of ASD PRS on the relationship between each covariate and *PEITANG* also was considered. At matched GWS thresholds, the polygenic risk for schizophrenia, educational attainment, and all six brain imaging phenotypes also maintained significant prediction of *PEITANG* after covarying for ASD (Table 2). The relationship between the polygenic risk for attention deficit hyperactivity disorder and *PEITANG* was not independent of the effects of ASD PRS.

**Figure 3.**
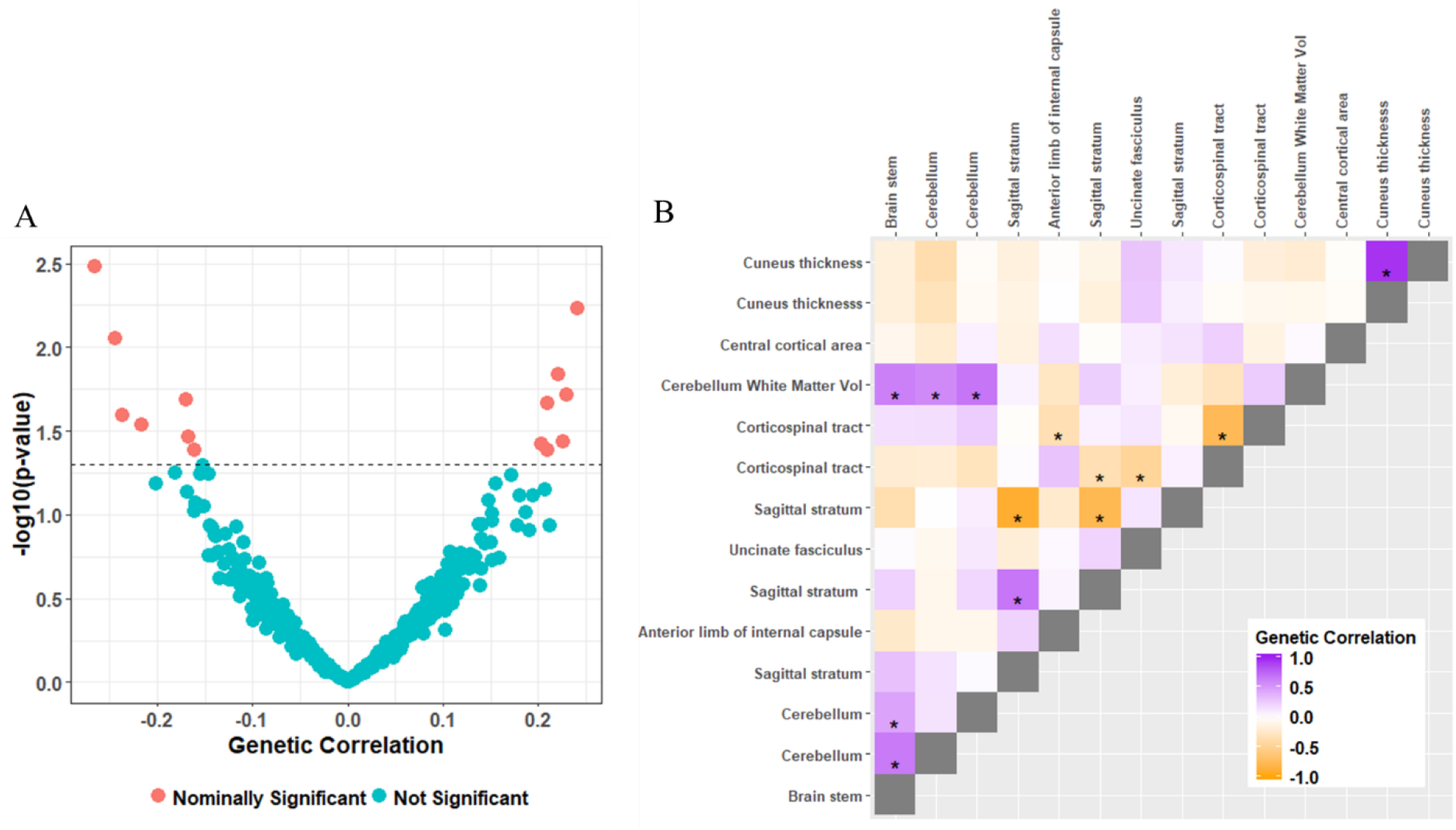
Selection of brain image-derived phenotypes as covariates in the polygenic risk score analyses between autism spectrum disorder (ASD) and neuropsychiatric traits in the young. (A) Genetic correlation between ASD 3,199 brain imaging phenotypes from the Brain Imaging Genetics project and (B) genetic correlation between 14 brain imaging phenotypes nominally genetically correlated with ASD.

**Figure 4.**
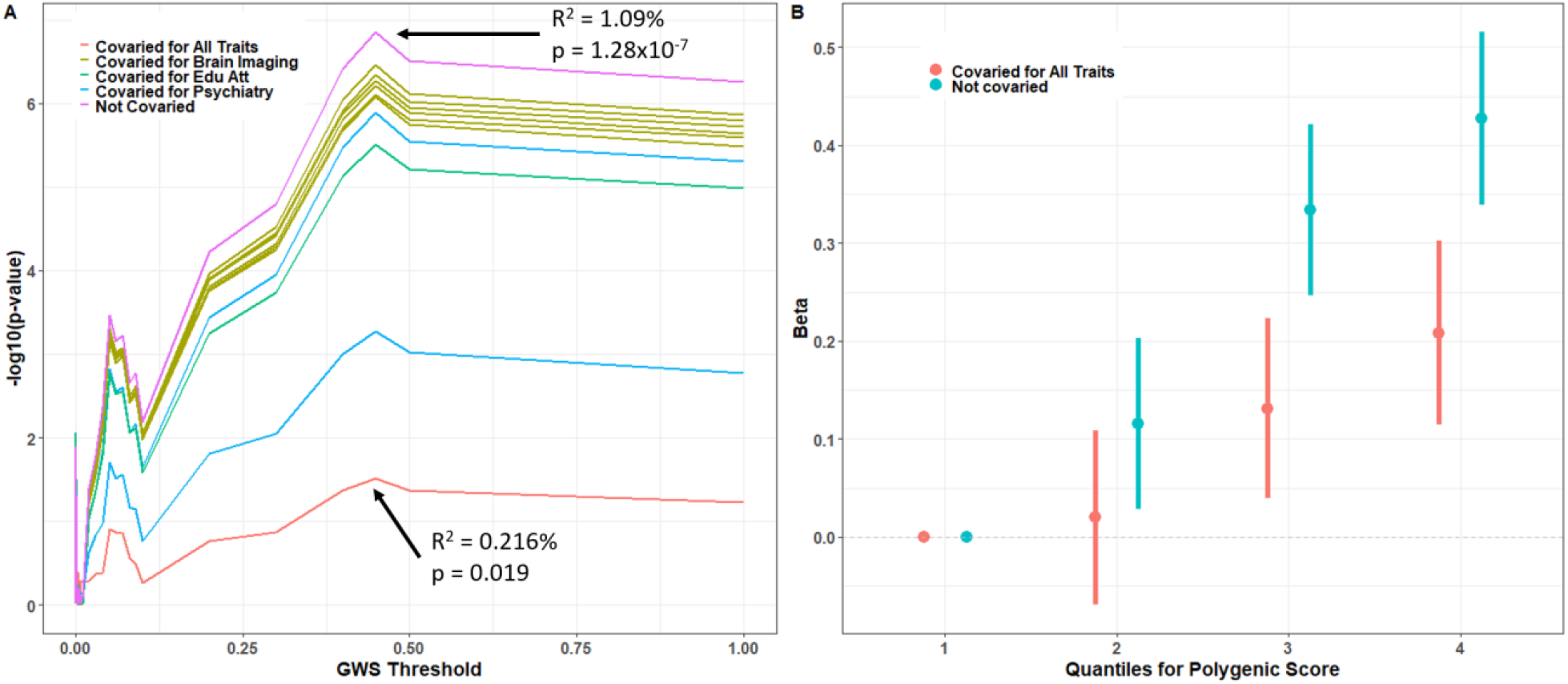
Prediction of ability to recognize angry faces in the middle proband group (ages 11-17) using the polygenic risk for autism spectrum disorder (ASD) covaried for age, sex, ten principle components, and the PRS for four brain imaging phenotypes, attention deficit hyperactivity disorder, schizophrenia, and educational attainment. (A) PRS p-value across a range of genome-wide significance (GWS) thresholds; the maximum PRS before and after covarying for all brain and psychiatry traits are labeled. (B) The positive correlation between quartiles of ASD polygenic risk and the number of correct responses to anger recognition trials in the Penn Emotional Intelligence Test; the lowest PRS quartile represents the referent.

**Figure 5.**
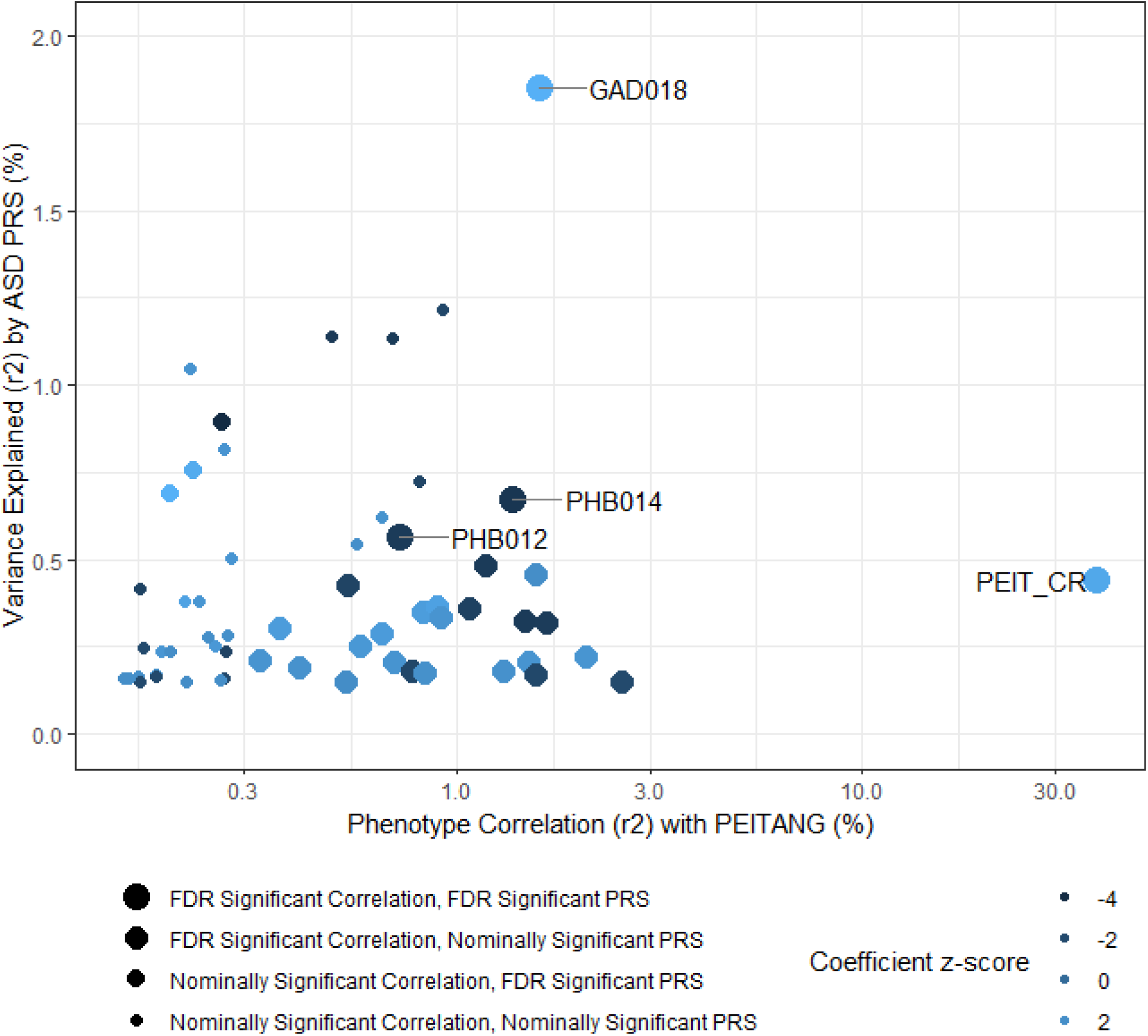
Fifty-nine traits phenotypically correlated with *PEITANG* and at least nominally predicted by polygenic risk (PRS) for autism spectrum disorder (ASD) in the Philadelphia Neurodevelopmental Cohort middle proband group (age 11-17). Neurodevelopmental traits are labeled if the correlation with *PEITANG* and PRS survive multiple testing correction; these are *PEIT_CR*: Penn Emotion Identification Test total correct responses for all trials; *GAD018*: Generalized Anxiety Disorder: Did you feel any of the following physical symptoms when you worried the most: irritability (feeling easily annoyed)?; *PHB012*: Specific Phobia: Thinking about all of the time that you were afraid of (insert worst fear), whether or not you actually faced it, how long did this fear last? (Days); *PHB014*: Specific Phobia: Thinking about all of the time that you were afraid of (insert worst fear), whether or not you actually faced it, how long did this fear last? (Months).

### Predicting Informant Perception

Self-reported behavioral attributes (i.e., the person’s perception of their own trait or behavior) may deviate from others’ perception of that same attribute.^22^ We use ASD polygenic risk scores to evaluate informant perception of traits in the middle proband group and the differences between perceived and self-reported phenotypes. After correction for multiple testing based on the number of phenotypes tested (N = 507; FDR Q < 0.05), 24 informant-reported phenotypes were significantly predicted by genetic liability to ASD. The most significantly predicted informant-reported phenotype was *PHB014: Specific Phobia: Thinking about all of the time that you were afraid of (insert worst fear), whether or not you actually faced it, how long did this fear last (Months)?* (informant-reported *PHB014*: z-score = −3.86, R^2^ = 1.20%, p = 1.18×10^−4^) and agreed with the proband-reported effect direction after covarying for *PEITANG* (proband-reported *PHB014*: z-score = −3.01, R^2^ = 0.156%, p = 0.001). The remaining 23 informant-reported phenotypes predicted by polygenic risk to ASD share general themes of perceived poor mood and poor behavior of the child by the informant (Table S5). Of these 23 informant-reported phenotypes, three of the proband-reported counterparts also were significantly predicted by ASD PRS after covarying for the effects of *PEITANG*, two of which had consistent effect direction with the informant (*SIP039: SIPS-Structured Interview for Prodromal Symptoms: Within the past 6 months, are you having a harder time getting normal activities done?* and *SCR006: General Probes: Are you currently taking medication because of your emotions and/or behaviors?*) and one with opposing effects directions relative to the informant (*PHB012: Specific Phobia: Thinking about all of the time that you were afraid of (insert worst fear), whether or not you actually faced it, how long did this fear last? (Days)*).

To evaluate differences in self-versus informant-perceived traits, 198 new phenotypes were created, each with at least 500 PNC probands: the differences between 11-to-17-year-old proband-reported behavior and 11-to-17-year-old informant-reported behavior. Polygenic risk for ASD predicted 41 differences in self-reported and perceived neuropsychiatric phenotypes (FDR Q < 0.05) which tend to highlight differences in outwardly visible traits such as attention difficulties, eating abnormalities, and defiance of authority figures (Table S6). The phenotype differences (see Methods) most strongly predicted by polygenic risk for ASD were *Structured Interview for Prodromal Symptoms (SIP) 039:* “*Within the past 6 months, are you having a harder time getting normal activities done?”* (z-score = 3.90, R^2^ = 0.591%, p=9.92×10^−5^) and *SIP025*: “*I think that I may hear my own thoughts being said out loud*” (z-score = 3.96, R^2^ = 0.717%, p = 7.77×10^−5^). The top 10% of phenotypes with classification differences represented a mean of 38.4% ± 3.82 of participants per phenotype classified differently by their informant, all of which were representative of trait designation differences (e.g., the proband answered “yes” while the informant answered “no”). Four phenotypes with classification differences were significantly predicted by ASD PRS after correction for multiple testing (FDR Q < 5%): *DEP010: Depression: During that time when you were feeling the most (sad, grouchy, irritable, in a bad mood, had trouble having fun), did that/those feeling(s) last most of the day?* (z-score = −2.73, R^2^ = 0.013%, p = 0.007), *GAD003A: Generalized Anxiety Disorder: Please look at this list and tell me if you worry a lot about: Your performance in school and/or sports* (z-score = - 3.06, R^2^ = 0.010%, p = 0.002), *GAD017: Generalized Anxiety Disorder: Did you feel any of the following physical symptoms when you worried the most: concentration problems (trouble focusing or paying attention)?* (z-score = 3.06, R^2^ = 0.018%, p = 0.002), *SOC008: Social Anxiety: Did this bother you more than most people your age?* (z-score = −2.95, R^2^ = 0.013%, p = 0.003).

## Discussion

### Accurate Recognition of Emotions

Polygenic risk for ASD predicted several aspects of facial emotion recognition in healthy subjects aged 11 to 17 years old. Thus, our results tie the genetic risk for ASD pathology directly to related traits in the normal population. We showed that ASD PRS was positively associated with the ability to correctly recognize angry faces (i.e., *PEITANG*) when given the option of angry, fearful, happy, neutral, or sad as delivered in PEIT. Negative facial emotion recognition deficits are typically considered hallmark attributes of ASD cases^1^ although this is often contested in the literature on the basis of different diagnostic instrument accuracy, ASD case severity heterogeneity, and eye movement tracking frequency and accuracy.^23, 24^ Notably, after covarying all other facial emotion recognition measure from PEIT on correctness in anger trials, polygenic risk for ASD also predicted poor recognition of happy and neutral faces in others. We speculate that this difference between angry and other facial emotions is linked to the increased ability to recognize the emotions most familiar to the child. ASD toddlers experience more intense anger than typically-developing (i.e., children not affected by ASD) and developmentally-delayed children (defined by Macari, *et al*. as children with global or specific language, communication, or motor delays)^25, 26^ but ASD severity was not associated with anger intensity in these studies. Furthermore, caring for a child with ASD has been linked to increased stress, anger, and salivary cortisol levels,^27^ providing additional mechanisms for prolonged anger emotion exposure and heightened anger emotion memory in ASD youths (because the ASD child may be evoking anger in a caregiver).

Animal data are convergent with human findings. In rats, the effects of early adversity through caregiver-offspring interaction persisted into adulthood and manifested as altered expression of the oxytocin receptor in the amygdala.^28^ Human oxytocin levels and the amygdala brain region are robustly associated with emotional intelligence and have been implicated in reduced attention to angry faces (in relation to depressive symptoms)^29^ and behavioral responses to threat-emotions.^30^ We provide support for these observations using genetic data by identifying that, after covarying for correct anger recognition, genetic risk for ASD predicted proband-reported feelings of irritability (i.e., *GAD018: Generalized Anxiety Disorder: Did you feel any of the following physical symptoms when you worried the most: irritability (feeling easily annoyed)?*) with a positive effect direction (i.e., increased polygenic risk for ASD predicted more self-reported irritability). Furthermore, the irritability phenotype was significantly positively correlated with *PEITANG* suggesting that similar mechanisms or brain regions may act in similar ways to modulate internalized anger versus perception of anger in others.

### Speed of Emotion Recognition

Reaction times to recognize emotional faces also were measured with PEIT in the PNC. We demonstrated that emotion recognition reaction time was significantly predicted by ASD polygenic risk, though the magnitude of this effect was less than that of *PEITANG*. In this study, ASD PRS predicted faster reaction times to anger and fearful faces and slower reaction time to happy facial emotions. This is less explored in the ASD behavioral literature; however, there is epidemiological evidence that ASD cases and typically-developing children both avoid ruminating over facial emotion recognition tasks.^31^ Though not evaluated here, this epidemiological observation suggests that the polygenic risk for ASD also may predict reaction times to recognize facial emotions in ASD cases. This process has been linked to timed release of cortisol following emotional task ques.^32^ Emotional intelligence capabilities may be dependent on the context in which they are required. For example, conversational context clues and caregiver behavior have demonstrated effects on emotional intelligence but are not effectively reproduced in tests for emotion recognition and differentiation.^28, 33, 34^ Faster response to angry and fearful faces is perhaps linked to induction of acute stress at times of social interaction in youth. This stress has been linked to earlier visual processing following external stimuli^35, 36^, which may be less present or less readily apparent when individuals are confronted with less socially welcoming emotions like those of people with happy or neutral faces which were not robustly predicted in this study.

### Differentiating Emotion Intensity

Contrary to PEIT, PEDT shows participants two faces with the same emotion and asks, for example, which face is “more afraid.” Deficits in this social cognition task may be influenced by other contextual information from routine conversation not adequately incorporated into emotion differentiation tasks (i.e., perception of body language,^37^ tone and pitch,^38^ peer group interaction (i.e., ensemble perception),^39^ and familiarity with the interacting peer(s))^40^ and not solely on the ability to recognize emotion or general neuropsychiatric features of the ASD condition. While ASD PRS predicted recognition of angry faces and the speed with which these faces were identified, it did not predict the ability to distinguish emotion severity differences between faces if shown more than one (as measured by PEDT). This is an importantly distinct task that is less well discussed in the ASD literature. Though not specifically evaluated here due to our emphasis on youths unaffected by ASD, the absence of relationships between ASD polygenic risk and measures of emotion severity differentiation perhaps emphasizes the importance of conversational and environmental context clues from interpersonal interactions in the perception of emotional severity by ASD cases.^32^

### Effects of Other Brain and Psychiatric Phenotypes

The relationship between PRS for other psychiatric disorders and brain imaging phenotypes also predicted angry emotion recognition. While the relationship between ADHD and angry face recognition was not independent of the effects of ASD, genetic risk for schizophrenia, educational attainment, and all six brain imaging phenotypes significantly predicted the ability to recognize angry faces in others correctly. Genetic risk for schizophrenia, educational attainment, cerebellar regions, the uncinate fasciculus, and cuneus were positively associated with recognizing angry faces, perhaps even further highlighting the role of oxytocin, stress, and anger on the brain.^41, 42^

After covarying for several additional brain phenotypes (schizophrenia, ADHD, educational attainment, and six brain imaging phenotypes), the genetic risk for ASD remained positively associated with the ability to correctly recognize angry faces. This observation suggests that although PRS for other traits (e.g. schizophrenia) independently predicted recognition of angry faces with greater magnitude than ASD PRS, the relationship between ASD PRS and the ability to correctly recognize anger in others is independent of the effects of highly genetically correlated traits with greater sample sizes.

### Informant-Perceived Traits

Finally, we evaluated the genetic overlap between ASD polygenic risk and informant-perceived traits in probands aged 11 to 17 years. There was a negative relationship between genetic risk for ASD and perceived duration of fear of a specific phobia (*PHB014*) which was consistent with the proband-reported phenotype. Notably, ASD PRS also predicted several phenotypes we derived describing the difference between informant- and proband-reported phenotypes. There was an overall lack of consistency in trait reporting with up to 38% of probands classified differently by their informant (compared to their self-classification) with respect to some PNC traits. This observation is not novel and reinforces the need for use of several informants, where available. A majority of PNC middle proband informants were the proband’s mothers; however, the setting in which behavior is observed may be very different between informant mothers and, for example, informant teachers.^43^

### Limitations

This study has two main limitations. First, the PNC evaluates broad aspects of human neurodevelopment and with respect to social cognition only investigates facial stimuli at the exclusion of gesture, vocalization, and complex social situations. Future work may investigate the relationship between ASD PRS and higher resolution measures of social cognition. Second, ASD cases were removed prior to polygenic risk scoring but the remaining proband groups likely contain disproportionate ratios of disorders genetically correlated with ASD which may bias the observed predictions in a direction that favors ASD PRS strongly predicting a PNC trait (i.e., false positives). Given the strength of the relationship between ASD PRS and *PEITANG*, this potential bias likely lies within the phenotypes predicted by ASD PRS with nominal significance (Figure 1). Even though these biases may influence the specific traits predicted by ASD PRS in each proband group, the strongest relationship (ASD PRS and *PEITANG*) was replicable across age groups.

### Summary

We demonstrated that polygenic risk for ASD is associated with several neuropsychiatric features of ASD-unaffected individuals: (1) the ability to correctly recognize angry faces correctly in healthy youths and (2) the speed with which anger is recognized. Genetic risk for ASD was not associated with the ability to differentiate the intensity of facial emotions. These results were observed in a cohort without enrichment for specific neuropsychiatric phenotypes and self- and informant-reported ASD cases were removed from our analyses.^44^ The data presented support further investigation of how ASD risk alleles relate to psychopathology in healthy and ASD patients. Considering epidemiological data regarding facial emotion recognition, future work may investigate how ASD polygenic risk predicts correct recognition and reaction time for emotion recognition and differentiation tasks in ASD cases.

## Materials and Methods

### Genetic Data

Genome-wide association study (GWAS) summary statistics for ASD (18,382 cases and 27,969 controls of European descent) were obtained from Grove, *et al*. 2019 and accessed via the Psychiatric Genomics Consortium (PGC) webpage (available at https://www.med.unc.edu/pgc/results-and-downloads). GWAS summary statistics for additional psychiatric disorders also were accessed through the PGC webpage. GWAS summary statistics for 3,144 brain image-derived phenotypes were obtained from the Oxford Brain Imaging Genetics project (BIG; available at http://big.stats.ox.ac.uk/).^11, 21^ The sample sizes for each of the traits/psychiatric disorders used in this study are provided in Table S2.

Neuropsychiatric and genotype data for 9,267 youths aged 8-21 were obtained from the PNC (*Neurodevelopmental Genomics: Trajectories of Complex Phenotypes* dataset (dbGAP phs000607.v3.p2)). Comprehensive details of the clinical and cognitive assessments have been reported previously.^6-9^ Briefly, the collaboration between the Children’s Hospital of Philadelphia (CHOP) and University of Pennsylvania deeply phenotyped youth participants visiting CHOP or an affiliated clinic for routine visit and volunteered to participate in genomic studies of complex pediatric disorders. The cohort is considered generally healthy and is not enriched for any specific disorder, behavior, or trait. Participants were excluded from the PNC study if they (1) exhibited anxiety to the point of inability to complete the battery administered, (2) had any medical condition that may impact cognitive battery measurements including blindness, seizures, head trauma, or central nervous system tumor, and (3) had any medical condition that may interfere with magnetic resonance imaging (MRI) scanning including metallic inserts, orthopedic circumstances, or pregnancy. Participants were given a computerized neurocognitive battery and a subsample underwent neuroimaging. Clinical testing for each participant included GOASSES and a psychopathology symptom and criterion-related assessment; the computerized neurocognitive battery administered to each participant provides measures of executive control functions, episodic memory, complex cognitive processing, social cognition, and sensorimotor and motor speed. For a complete list of neurobehavioral domains assessed, representing approximately 900 phenotypes per participant, see https://www.ncbi.nlm.nih.gov/projects/gap/cgi-bin/study.cgi?study_id=phs000607.v3.p2.

The PNC consists of three proband groups classified by age (Table 1) and includes information from a second visit of the middle proband age group. Phenotypes were included in our analysis if they had ≥ 500 participants and effective sample size ≥ 50 (i.e., the number of cases necessary to detect an effect weighted by the proportion of the binary categories). To compare differences in informant-perceived versus self-reported phenotype, binary phenotypes with yes/no responses were recoded as 1 (proband-reported and informant-reported “yes”), 2 (self-reported “yes” and informant-reported “no”), 3 (self-reported “no” and informant-reported “yes”), and 4 (self-reported and informant-reported “no”). For quantitative traits, the new phenotype represented the difference between proband-reported and informant-reported values (i.e., proband value minus informant value). Ordinal phenotypes were recoded as 1 (more severe rating by proband), 2 (proband and informant reported same rating), and 3 (more sever rating by informant).

Probands were excluded from this study if they or their informant answered “yes” to *MED291: “Autism or Pervasive Developmental Disorder - Do/did you have this problem?”* Based on answers to MED291, fewer than 5% of each proband group was removed from PRS analyses.^44^

### GWAS Quality Control

Preimputation quality control was performed in plink v1.9; briefly: (1) SNPs were removed with call rate < 0.95; (2) Samples were removed with call rates < 0.95 and absolute value inbreeding coefficients > 0.2, (3) SNPs were removed with call rates < 0.98 and HWE p-values < 1×10-6. Individuals of European descent were confirmed on the basis of the genetic information via principle component analysis using SNPs shared across all beadchips (N = 38,739 SNPs) and the 1000 Genomes Project reference panel for populations with European ancestry (N=503). Cryptic relatedness among sample pairs was determined using a PI-HAT threshold of 0.2. For sample pairs with PI-HAT > 0.2, the sample with more informative phenotypes was retained. Imputation was performed for 4,309 unrelated individuals of European ancestry using SHAPEIT for pre-phasing,^45^ IMPUTE2 for imputation,^46^ and the human 1000 Genomes Project Phase 3 as a reference panel. Samples were stratified by proband group assignment in the PNC for PRS analyses (Table 1).

### SNP-Heritability and Genetic Correlation

SNP-based observed-scale heritability and genetic correlation for PGC psychiatric disorders and BIG brain imaging phenotypes were determined using the Linkage Disequilibrium Score Regression (LDSC) method.^20^

### Polygenic Risk Scoring

PRS based on ASD GWAS summary statistics were used to predict phenotypes in adult, middle, and young proband groups from the PNC. Polygenic risk scoring was performed in PRSice v2^10^ with the default settings (i.e., SNPs were clumped based on 250kb windows on either side of target SNP based on clump-r^2^ and clump-p thresholds of 0.1 and 1, respectively. All PRS were covaried for age, sex, and the first ten principle components (PCs). PRS predictions also were covaried using the PRS of UKB BIG brain imaging phenotypes and other psychiatric disorders (Table S2) to detect whether the signals between ASD and PNC traits were independent of the effects of additional psychiatric disorders, brain imaging phenotypes, and educational attainment. The individual genetic liabilities to ASD for PNC participants were binned into four quartiles and regressed against the dbGAP phenotype of interest using a generalized linear model including the same covariates as the respective PRS prediction (i.e., age, sex, 10 PCs, and PRS of brain imaging phenotypes and psychiatric phenotypes where appropriate). We applied a FDR multiple testing correction (FDR 5%) accounting for the number of PRS thresholds tested and the number of phenotypes investigated (N = 2,946,000 tests). Additionally, we considered as a suggestive significance a FDR multiple testing correction accounting for the number of phenotypes tested only (N = 491).

### Phenotype Correlation

Spearman’s correlation between phenotype measurements of the PNC were calculated in R studio using the rcorr function of the Hmisc library. Correlation p-values were adjusted for the number of tests performed using FDR Q < 0.05.

## Data Availability

All data used in this study are either publicly available summary data or available for any research to submit an application through dbGAP.

## Acknowledgements

This study was supported by the Simons Foundation Autism Research Initiative (SFARI Explorer Award: 534858) and the American Foundation for Suicide Prevention (YIG-1-109-16). CMC was supported by a Fundação de Amparo à Pesquisa do Estado de São Paulo (FAPESP 2018/05995-4) international fellowship. Support for the collection of the data for Philadelphia Neurodevelopment Cohort (PNC) was provided by grant RC2MH089983 awarded to Raquel Gur and RC2MH089924 awarded to Hakon Hakonarson. Subjects were recruited and genotyped through the Center for Applied Genomics (CAG) at The Children’s Hospital in Philadelphia (CHOP). Phenotypic data collection occurred at the CAG/CHOP and at the Brain Behavior Laboratory, University of Pennsylvania.

## Supplementary Figures

**Figure S1.**
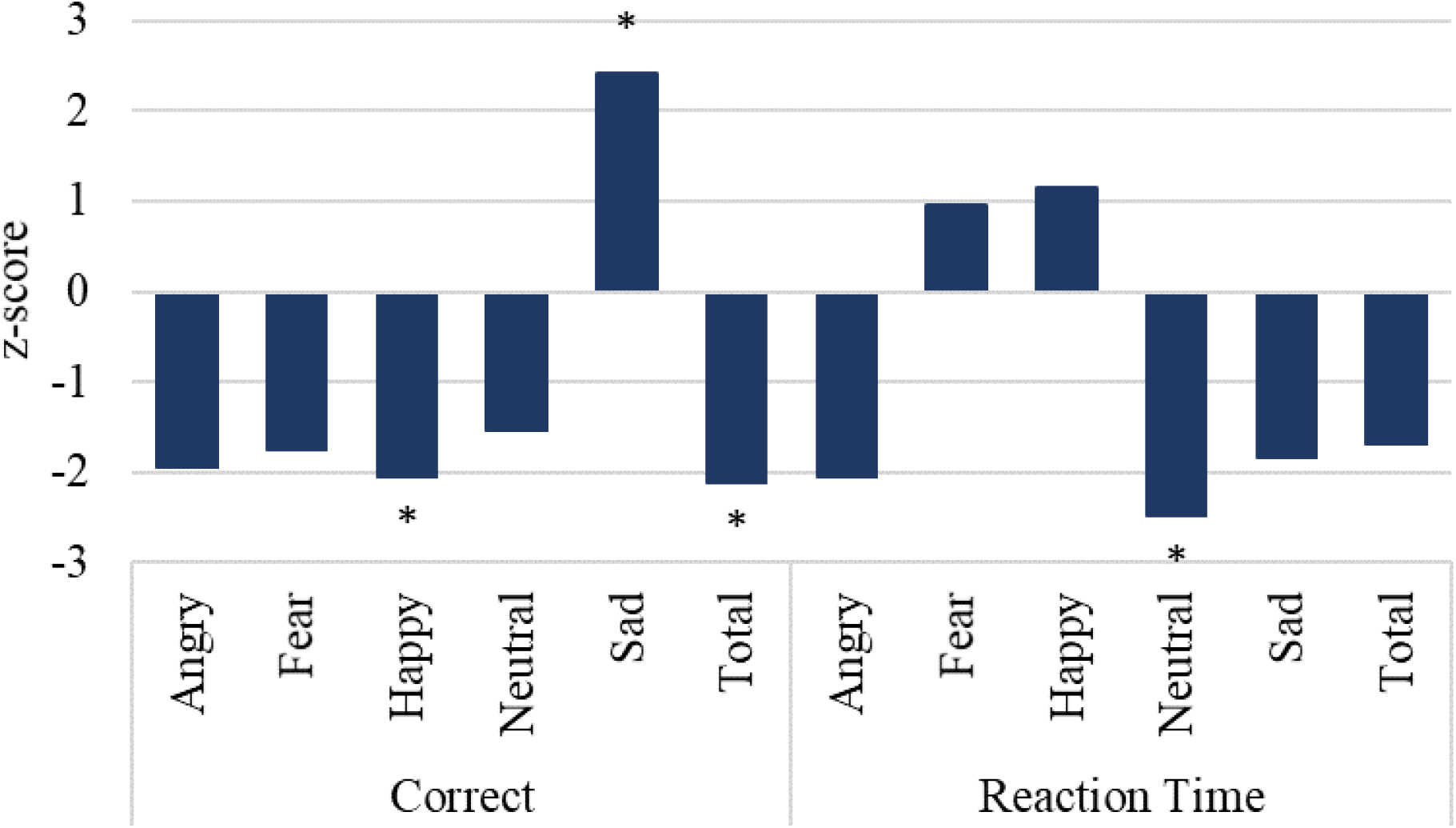
Prediction of facial emotion differentiation capabilities of the middle proband group (ages 11 to 17) of the Philadelphia Neurodevelopmental Cohort using polygenic risk (PRS) for autism spectrum disorder. The x-axis shows correctness and response time for each facial emotion using two neurocognitive instruments: The Penn Emotion Differentiation Test. Significance is indicated by * for p < 0.05.

**Figure S2.**
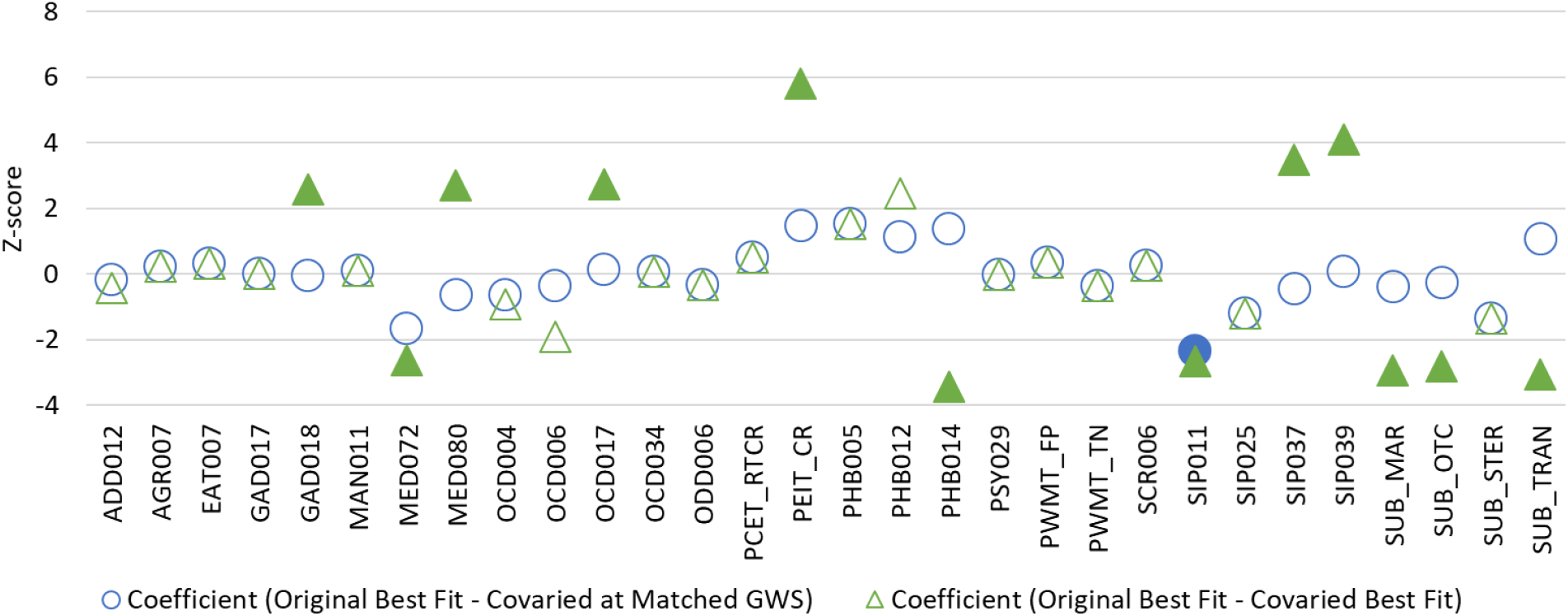
Z-score converted difference in PRS coefficient for 30 phenotypes suggestively predicted by polygenic risk for autism spectrum disorder in the middle proband group of the Philadelphia Neurodevelopmental Cohort. Two differences were evaluated: (1; circles) the coefficient difference between original best-fit PRS and the same PRS covaried for the effects of *PEITANG* (i.e., the same genome-wide significance threshold in both instances) and (2; triangles) the coefficient difference between original best-fit PRS and the best-fit PRS after covarying for the effects of *PEITANG* (i.e., the GWS may be different in each instance). Note that in several instances the covaried best-fit PRS is the same as the matched PRS from the original test. Filled-in shapes indicate statistically significant differences between original and covaried PRS coefficients (FDR Q < 0.05).

## Supplementary Tables

**Table S1.**
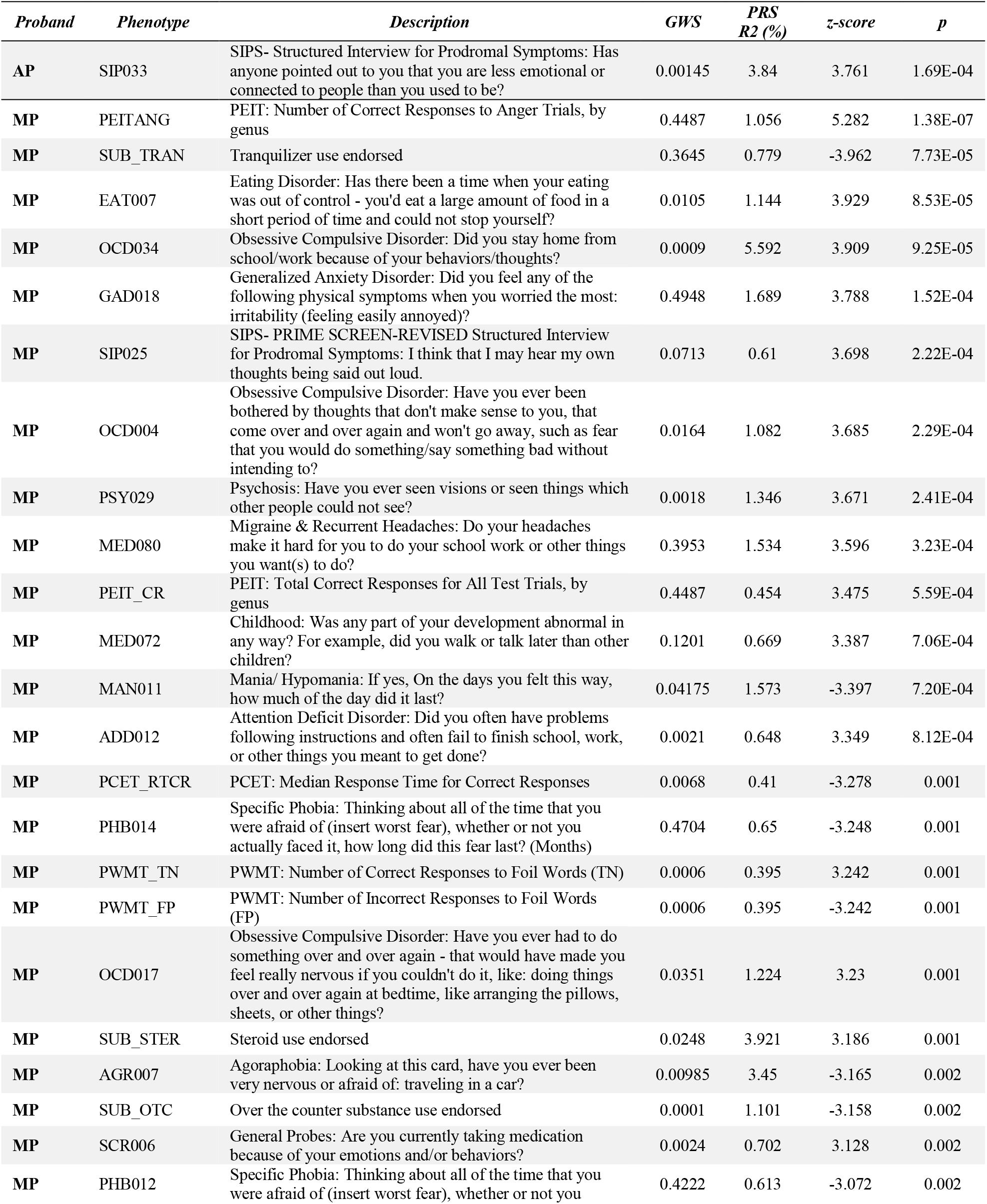
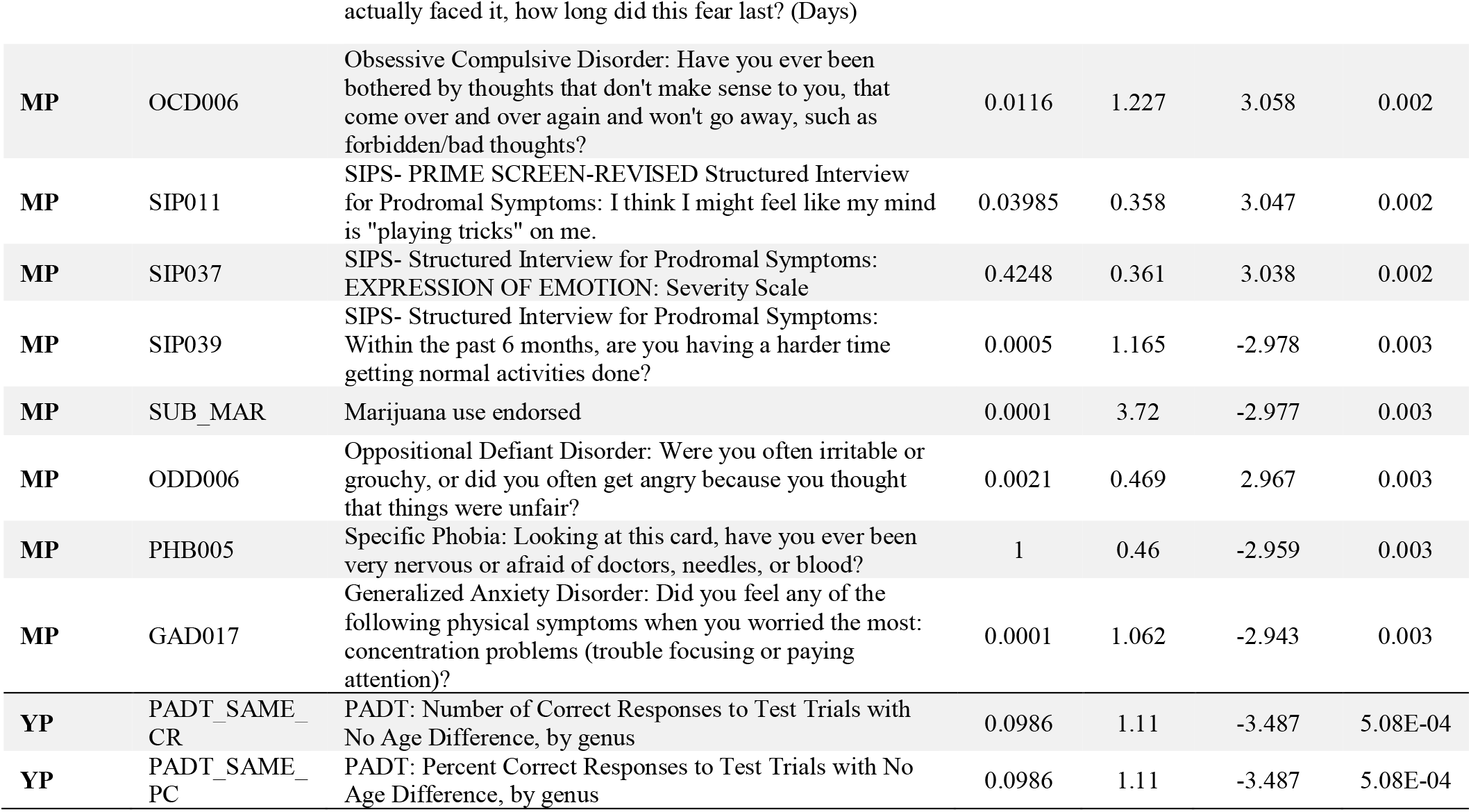
Phenotypes meeting suggestive threshold for significant prediction by polygenic risk for autism spectrum disorder in the young (YP), middle (MP), and adult (AP) proband groups.

**Table S2.** Correlation between PEITANG and other measures recorded by the Penn Emotion Identification Test (PEIT).

| **Trait** | **Emotion** | **Measure** | **r<sup>2</sup>** | **p** |
| --- | --- | --- | --- | --- |
| PEIT_CR | All | Correct | 0.378 | 2.20E-16 |
| PEIT_CRT | All | Reaction Time | 0.005 | 3.37E-04 |
| PEITANGRT | Anger | Reaction Time | 0.012 | 2.87E-08 |
| PEITFEAR | Fear | Correct | 0.007 | 1.86E-05 |
| PEITFEARRT | Fear | Reaction Time | 0.002 | 3.79E-02 |
| PEITHAP | Happiness | Correct | 0.022 | 1.85E-14 |
| PEITHAPRT | Happiness | Reaction Time | 0.008 | 2.96E-06 |
| PEITNOE | Neutral | Correct | 0.008 | 6.39E-06 |
| PEITNOERT | Neutral | Reaction Time | 0.001 | 8.00E-02 |
| PEITSAD | Sad | Correct | 0.20 | 7.46E-13 |
| PEITSADRT | Sad | Reaction Time | 0.003 | 1.02E-02 |

**Table S3.**
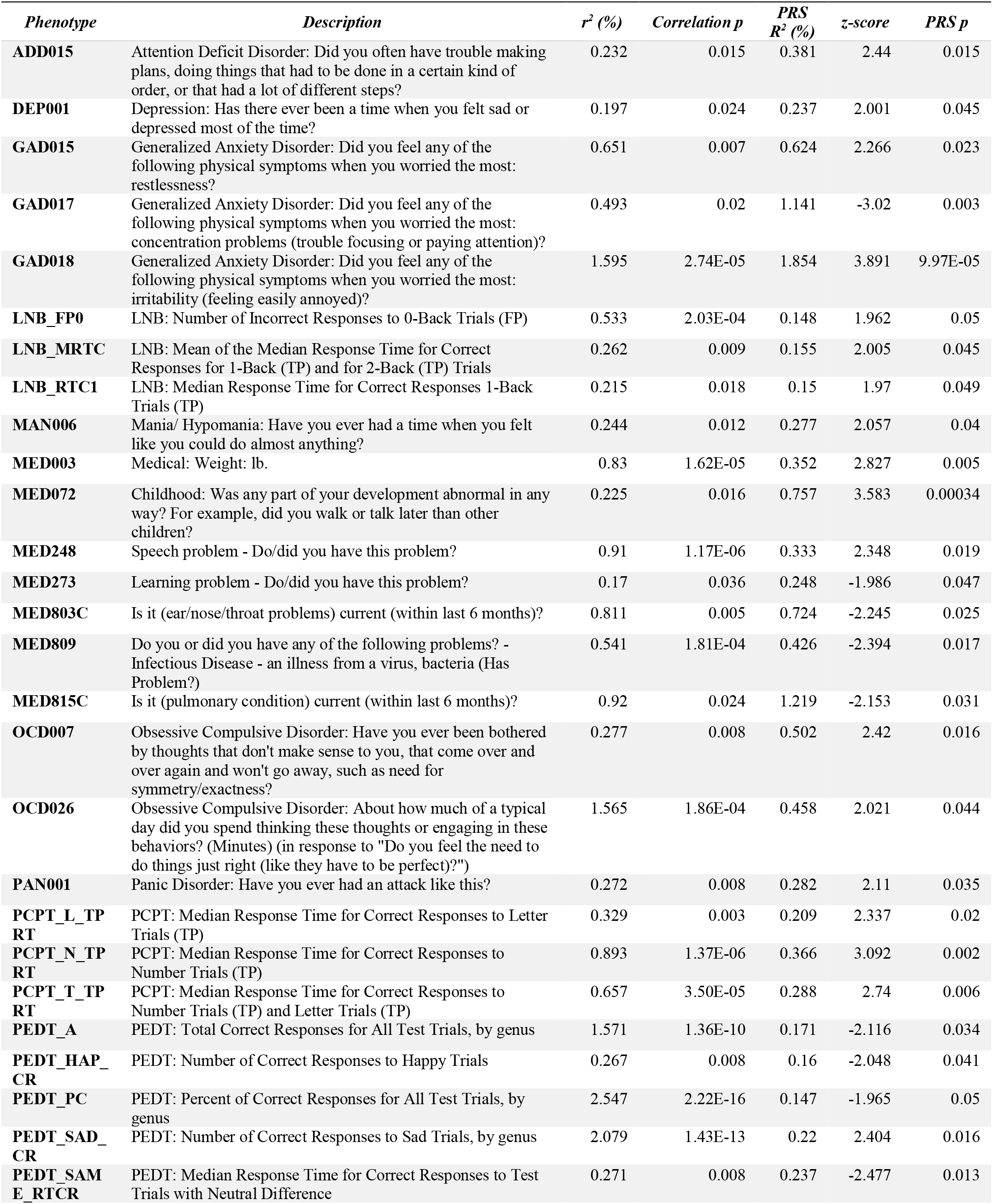
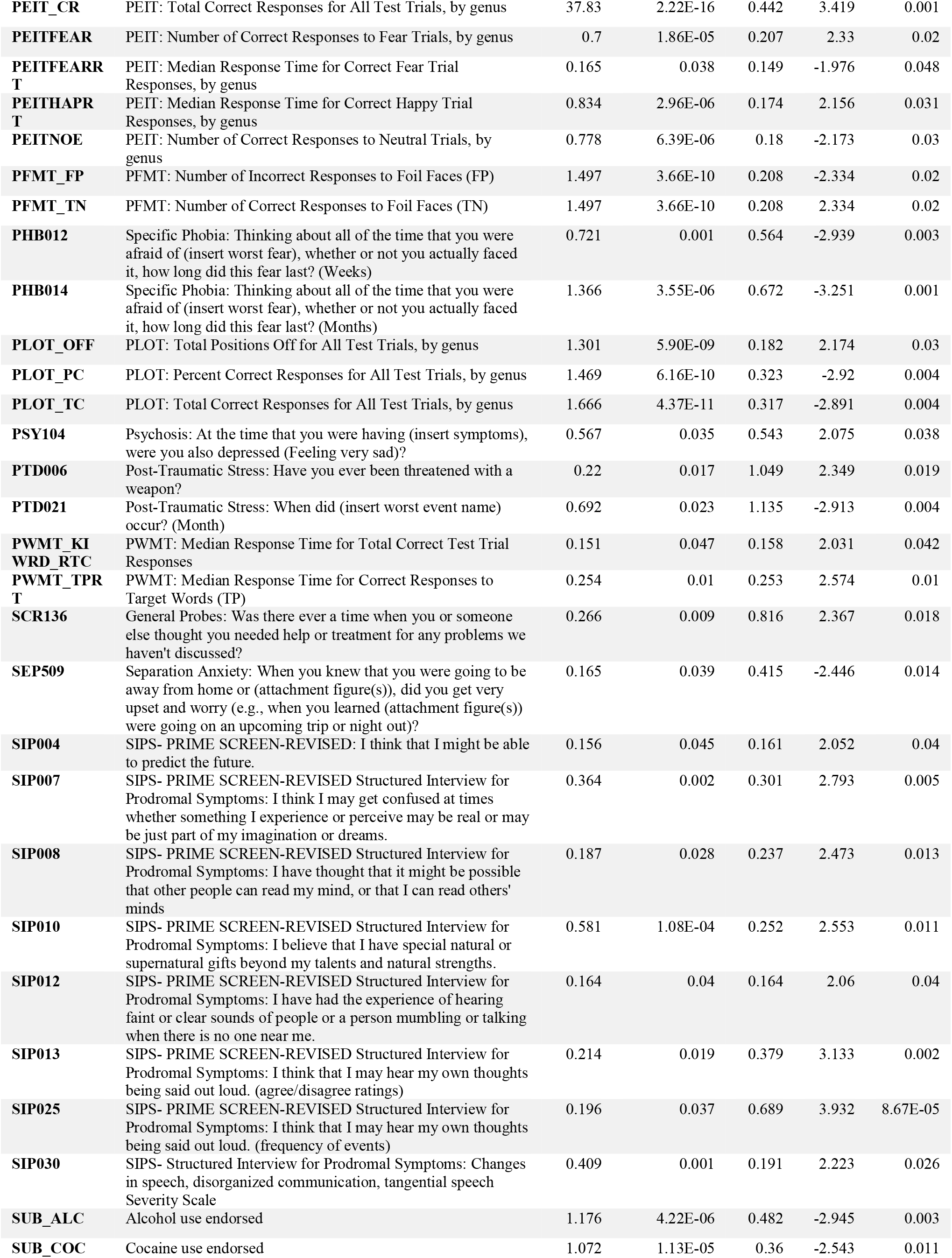
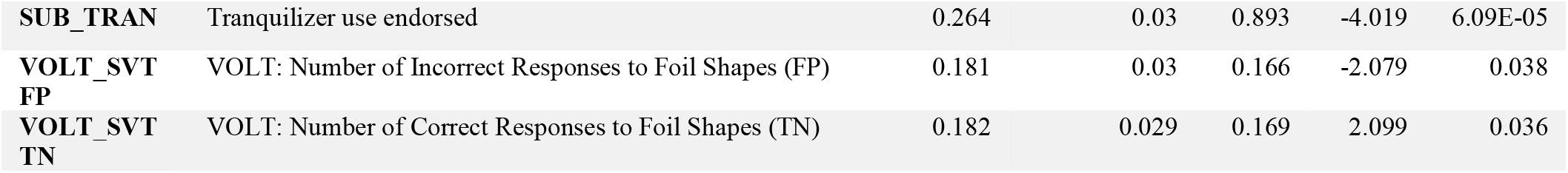
Traits significantly correlated with PEITANG in the middle proband group of the Philadelphia Neurodevelopmental Cohort and at least nominally predicted by polygenic risk (PRS) for autism spectrum disorder.

**Table S4.**
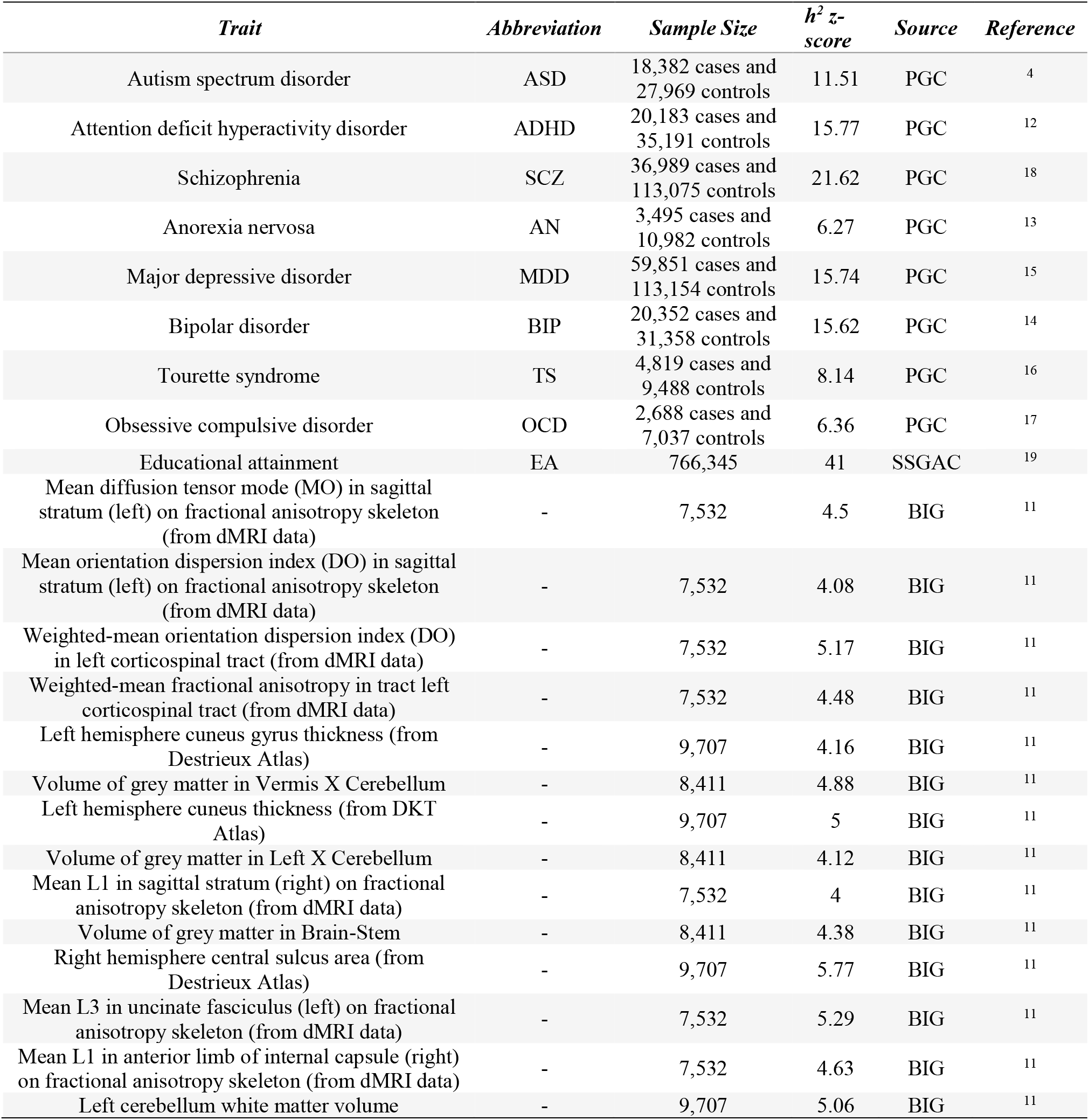
Samples size and SNP-based observed-scale heritability (h^2^) for genome-wide association study summary statistics used in this study.

**Table S5.** Phenotypes predicted by polygenic risk for autism spectrum disorder based on perceived behavior from middle age group (11 to 17 years old) informants of the Philadelphia Neurodevelopmental cohort.

| *Phenotype* | *Description* | *GWS*<br>*Threshold* | *R2*<br>*(%)* | *z-*<br>*score* | *p* | *FDR*<br>*Q* |
| --- | --- | --- | --- | --- | --- | --- |
| **ADD020** | Did you often have difficulty sitting still for more than a few minutes at a time, even after being asked to stay seated, or did you often fidget with your hands or feet or wiggle in your seat or were you "always on the go"? | 0.0152 | 0.633 | 3.27 | 1.17E-04 | 0.037 |
| **ADD028** | Did family members seem upset, angry, or annoyed with you because of your difficulties? | 0.326 | 1.62 | 3.60 | 0.00 | 0.023 |
| **ADD029** | Did these behaviors/inattention bother your friends? | 0.2256 | 1.66 | 3.62 | 3.39E-04 | 0.023 |
| **AGR003** | Agoraphobia: Looking at this card, have you ever been very nervous or afraid of: being in an open field? | 0.00145 | 8.57 | 3.15 | 3.19E-04 | 0.045 |
| **CDD026** | Conduct Disorder: How old were you the first time you did these (list behaviors)? (Age) | 0.0141 | 3.66 | -3.13 | 0.002 | 0.045 |
| **DEP009** | When did you feel the most (sad, grouchy, irritable, in a bad mood, had trouble having fun)? | 0.00195 | 1.12 | 3.27 | 0.002 | 0.037 |
| **DEP014** | Depression: During this time, did you have trouble sitting still or feel like you had to keep moving around OR did you move or think more slowly than usual? | 0.0006 | 3.63 | -3.26 | 0.001 | 0.037 |
| **DEP017** | Depression: During this time, did you blame yourself for bad things that happened or feel like you didn't really matter? | 0.406 | 3.28 | 3.10 | 0.001 | 0.045 |
| **MAN009** | Mania/ Hypomania: Was this different from how you usually are? (Follow up to "Has there ever been a time when you felt unusually grouchy, cranky, or irritable; when the smallest things would make you really mad?" | 0.4759 | 2.69 | 3.32 | 0.002 | 0.037 |
| **MAN026** | Mania/ Hypomania: During this time when you felt the most (too happy/excited/grouchy/energetic) did you become more interested in sex or more sexually active? | 0.01295 | 9.41 | -3.62 | 9.04E-04 | 0.023 |
| **ODD025** | Oppositional Defiant Disorder: How old were you the first time you did these? (Age) (in response to Did you stay home or were you sent home from school/work because of your behavior?) | 0.1177 | 1.28 | -3.53 | 2.95E-04 | 0.025 |
| **PAN012** | Panic Disorder: How much did having these [panic] attacks upset or bother you? | 0.05175 | 18.27 | 3.28 | 4.35E-04 | 0.045 |
| **PHB012** | Specific Phobia: Thinking about all of the time that you were afraid of (insert worst fear), whether or not you actually faced it, how long did this fear last? (Days) | 0.0008 | 1.14 | 3.77 | 0.002 | 0.023 |
| **PHB014** | Specific Phobia: Thinking about all of the time that you were afraid of (insert worst fear), whether or not you actually faced it, how long did this fear last? (Months) | 0.014 | 1.20 | -3.86 | 1.73E-04 | 0.023 |
| **PHB024A** | Specific Phobia: How old were you the first time you had this fear of (insert worst fear)? (Age) | 0.3757 | 2.00 | -3.14 | 0.002 | 0.045 |
| **PSY003** | Psychosis: How many voices did you hear? | 0.00015 | 19.40 | -3.57 | 7.38E-04 | 0.037 |
| **PSY017** | Psychosis: How long was the longest time that this [hearing voices] lasted? (Estimate duration; can be intermittent over this time) (Years) | 0.0002 | 20.70 | -3.79 | 3.58E-04 | 0.023 |
| **PSY054** | Psychosis: How many times have you smelled strange odors like this that other people couldn't smell? | 0.00155 | 48.68 | 4.48 | 2.07E-04 | 0.023 |
| **PTD007** | Post-Traumatic Stress: Have you ever been in a bad accident? | 0.0592 | 1.23 | -3.67 | 2.45E-04 | 0.023 |
| **PTD009** | Post-Traumatic Stress: Have you ever been very upset by seeing a dead body or by seeing pictures of the dead body of somebody you knew well? | 0.0019 | 0.809 | 3.14 | 0.002 | 0.045 |
| **SCR004** | General Probes: How long did you see someone [a counselor] in total? | 0.00825 | 1.43 | 3.28 | 0.001 | 0.037 |
| **SCR006** | General Probes: Are you currently taking medication because of your emotions and/or behaviors? | 0.3954 | 0.667 | 3.06 | 0.002 | 0.048 |
| **SIP036** | SIPS- Structured Interview for Prodromal Symptoms: Changes in perception of self, others, or the world in general: How long has it been since you first had this experience? | 0.01055 | 46.96 | -3.98 | 0.001 | 0.042 |
| **SIP039** | SIPS- Structured Interview for Prodromal Symptoms: Within the past 6 months, are you having a harder time getting normal activities done? | 0.0006 | 1.07 | -3.36 | 7.77E-04 | 0.037 |

**Table S6.** Phenotypes predicted by polygenic risk for autism spectrum disorder based on the difference between self-reported and perceived behavior from the middle age group (11 to 17 years old) of the Philadelphia Neurodevelopmental Cohort.

| *Phenotype* | *Description* | *GWS Threshold* | *R2 (%)* | *z-score* | *p* | *FDR Q* |
| --- | --- | --- | --- | --- | --- | --- |
| **ADD011** | Attention Deficit Disorder: Did you often have trouble paying attention or keeping your mind on your school, work, chores, or other activities that you were doing? | 0.00255 | 0.300 | -2.783 | 0.005 | 0.040 |
| **ADD012** | Attention Deficit Disorder: Did you often have problems following instructions and often fail to finish school, work, or other things you meant to get done? | 0.0019 | 0.445 | -3.404 | 6.74E-04 | 0.026 |
| **ADD014** | Attention Deficit Disorder: Did you often lose things you needed for school or projects at home (assignments or books) or make careless mistakes in school work or other activities? | 0.001 | 0.257 | -2.579 | 0.010 | 0.048 |
| **ADD015** | Attention Deficit Disorder: Did you often have trouble making plans, doing things that had to be done in a certain kind of order, or that had a lot of different steps? | 0.0018 | 0.287 | -2.717 | 0.007 | 0.040 |
| **AGR007** | Agoraphobia: Looking at this card, have you ever been very nervous or afraid of: traveling in a car? | 0.00985 | 0.362 | 3.049 | 0.002 | 0.029 |
| **CDD001** | Conduct Disorder: Was there ever a time when you often did things that got you into trouble with adults like lying or stealing (something worth more than \$5, from family, others, or stores)? | 0.2612 | 0.275 | 2.679 | 0.007 | 0.043 |
| **CDD007** | Conduct Disorder: Did you ever: try to hurt someone with a weapon (a bat, brick, broken bottle, knife, or gun)? | 0.00315 | 0.267 | -2.629 | 0.009 | 0.046 |
| **DEP010** | Depression: During that time when you were feeling the most (sad, grouchy, irritable, in a bad mood, had trouble having fun), did that/those feeling(s) last most of the day? | 0.04685 | 1.307 | -2.726 | 0.007 | 0.040 |
| **EAT007** | Eating Disorder: Has there been a time when your eating was out of control - you'd eat a large amount of food in a short period of time and could not stop yourself? | 0.0134 | 0.518 | -3.644 | 2.74E-04 | 0.013 |
| **GAD003 A** | Generalized Anxiety Disorder: Please look at this list and tell me if you worry a lot about: Your performance in school and/or sports | 0.01575 | 0.968 | -3.064 | 0.002 | 0.029 |
| **GAD012** | Generalized Anxiety Disorder: (If GAD011 =yes) Was it hard for you to stop yourself from worrying about these things (endorsed in before/after q's)? | 0.1727 | 1.360 | -2.623 | 0.009 | 0.046 |
| **GAD017** | Generalized Anxiety Disorder: Did you feel any of the following physical symptoms when you worried the most: concentration problems (trouble focusing or paying attention)? | 1.00E-04 | 1.770 | 3.059 | 0.002 | 0.029 |
| **MAN005** | Mania/ Hypomania: Have you ever had a time when you felt much more happy or excited than you usually do when there was nothing special going on? | 0.00315 | 0.263 | 2.598 | 0.009 | 0.046 |
| **OCD004** | Obsessive Compulsive Disorder: Have you ever been bothered by thoughts that don't make sense to you, that come over and over again and won't go away, such as fear that you would do something/say something bad without intending to? | 0.0185 | 0.550 | -3.758 | 1.75E-04 | 0.011 |
| **OCD006** | Obsessive Compulsive Disorder: Have you ever been bothered by thoughts that don't make sense to you, that come over and over again and won't go away, such as forbidden/bad thoughts? | 0.0116 | 0.372 | -3.068 | 0.002 | 0.029 |
| **OCD007** | Obsessive Compulsive Disorder: Have you ever been bothered by thoughts that don't make sense to you, that come over and over again and won't go away, such as need for symmetry/exactness? | 0.0306 | 0.290 | -2.716 | 0.007 | 0.040 |
| **OCD012** | Obsessive Compulsive Disorder: Have you ever had to do something over and over again - that would have made you feel really nervous if you couldn't do it, like: counting? | 0.0913 | 0.320 | -2.862 | 0.004 | 0.040 |
| **OCD016** | Obsessive Compulsive Disorder: Have you ever had to do something over and over again - that would have made you feel really nervous if you couldn't do it, like: ordering or arranging things? | 0.01065 | 0.403 | -3.231 | 0.001 | 0.029 |
| **OCD017** | Obsessive Compulsive Disorder: Have you ever had to do something over and over again - that would have made you feel really nervous if you couldn't do it, like: doing things over and over again at bedtime, like arranging the pillows, sheets, or other things? | 0.0355 | 0.306 | -2.801 | 0.005 | 0.040 |
| **ODD006** | Oppositional Defiant Disorder: Were you often irritable or grouchy, or did you often get angry because you thought that things were unfair? | 0.0021 | 0.310 | -2.831 | 0.005 | 0.040 |
| **ODD018** | Oppositional Defiant Disorder: You told me that you (list endorsed behaviors). How much did having these behaviors upset or bother you? | 0.00015 | 1.262 | -3.065 | 0.002 | 0.029 |
| **ODD020** | Oppositional Defiant Disorder: How much did these behaviors change your relationships with your friends? (Did you find yourself spending much less time than usual with your friends? Did your friends say something about your behavior?) | 0.1041 | 1.181 | -2.948 | 0.003 | 0.034 |
| **ODD021** | Oppositional Defiant Disorder: How much did these behaviors change your relationships with your teachers or classmates (co-workers/supervisors)? change how well you did at school, on tests, homework, or grades (or at job duties)? (in response to "were you often irritable or grouchy, or did you often get angry because you thought that things were unfair?") | 0.1326 | 1.292 | -3.103 | 0.002 | 0.029 |
| **PHB008** | Specific Phobia: Looking at this card, have you ever been very nervous or afraid of any other things or situations? | 3.00E-04 | 0.267 | -2.599 | 0.009 | 0.046 |
| **PHB012** | Specific Phobia: Thinking about all of the time that you were afraid of (insert worst fear), whether or not you actually faced it, how long did this fear last? (Days) | 0.4222 | 0.659 | 2.659 | 0.008 | 0.045 |
| **PSY029** | Psychosis: Have you ever seen visions or seen things which other people could not see? | 0.0017 | 0.316 | -2.842 | 0.004 | 0.040 |
| **PTD009** | Post-Traumatic Stress: Have you ever been very upset by seeing a dead body or by seeing pictures of the dead body of somebody you knew well? | 0.0026 | 0.415 | -3.258 | 0.001 | 0.029 |
| **SCR006** | General Probes: Are you currently taking medication because of your emotions and/or behaviors? | 0.0024 | 0.389 | -3.171 | 0.001 | 0.029 |
| **SEP512** | Separation Anxiety: How long was your longest period of worry about your (attachment figures)? (Days) | 0.00025 | 1.346 | 2.806 | 0.005 | 0.040 |
| **SEP514** | Separation Anxiety: How long was your longest period of worry about your (attachment figures)? (Months) | 0.00135 | 1.159 | -2.614 | 0.009 | 0.046 |
| **SIP004** | SIPS- PRIME SCREEN-REVISED: I think that I might be able to predict the future. | 0.0052 | 0.287 | 2.73 | 0.006 | 0.040 |
| **SIP007** | SIPS- PRIME SCREEN-REVISED Structured Interview for Prodromal Symptoms: I think I may get confused at times whether something I experience or perceive may be real or may be just part of my imagination or dreams. | 0.0032 | 0.282 | 2.714 | 0.007 | 0.040 |
| **SIP010** | SIPS- PRIME SCREEN-REVISED Structured Interview for Prodromal Symptoms: I believe that I have special natural or supernatural gifts beyond my talents and natural strengths. | 2.00E-04 | 0.367 | 3.097 | 0.002 | 0.029 |
| **SIP012** | SIPS- PRIME SCREEN-REVISED Structured Interview for Prodromal Symptoms: I have had the experience of hearing faint or clear sounds of people or a person mumbling or talking when there is no one near me. | 0.00175 | 0.323 | 2.9 | 0.004 | 0.037 |
| **SIP013** | SIPS- PRIME SCREEN-REVISED Structured Interview for Prodromal Symptoms: I think that I may hear my own thoughts being said out loud. | 0.01045 | 0.290 | 2.739 | 0.006 | 0.040 |
| **SIP025** | SIPS- PRIME SCREEN-REVISED Structured Interview for Prodromal Symptoms: I think that I may hear my own thoughts being said out loud. (only completed if participant agreed to SIP013) | 0.0713 | 0.717 | 3.959 | 7.77E-05 | 0.010 |
| **SIP037** | SIPS- Structured Interview for Prodromal Symptoms: EXPRESSION OF EMOTION: Severity Scale | 0.3765 | 0.308 | 2.709 | 0.007 | 0.040 |
| **SIP039** | SIPS- Structured Interview for Prodromal Symptoms: Within the past 6 months, are you having a harder time getting normal activities done? | 0.00055 | 0.591 | 3.899 | 9.92E-05 | 0.010 |
| **SOC002** | Social Anxiety: Looking at this card, was there ever a time in your life when you felt afraid or uncomfortable talking on the telephone or with people your own age who you don't know very well? | 0.01335 | 0.370 | -3.073 | 0.002 | 0.029 |
| **SOC008** | Social Anxiety: Did this bother you more than most people your age? | 0.0484 | 1.28 | -2.949 | 0.003 | 0.034 |
| **SOC011** | Social Anxiety: Thinking about all of the time that you were afraid of (insert worst fear), whether or not you actually faced it, how long did your fear of this situation last? (Months) | 0.3835 | 1.65 | 3.034 | 0.002 | 0.029 |

